# Prevalence of cardiometabolic risk factors according to urbanization level, gender and age, in apparently healthy adults living in Gabon, Central Africa

**DOI:** 10.1101/2023.05.04.23289536

**Authors:** Mérédith Flore Ada Mengome, Héléna Néoline Kono, Elsa Ayo Bivigou, Noé Patrick M’Bondoukwe, Jacques-Mari Ndong Ngomo, Bridy Moutombi Ditombi, Bedrich Pongui Ngondza, Cyrille Bisseye, Denise Patricia Mawili-Mboumba, Marielle Karine Bouyou Akotet

## Abstract

The prevalence of cardiometabolic risk factors (CMRFs) is increasing in sub-Saharan Africa and represents a serious health issue. Specific and accurate data are required to implement prevention programs and healthcare strategies. Thus, the aim of this study was to estimate the prevalence rates of CMRFs according to the level of urbanization, age and gender in Gabon.

A cross-sectional study was conducted using the World Health Organization’s (WHO) stepwise approach for the surveillance of chronic disease risk factors. Participants over 18 years of age, without known underlying disease, from rural and urban areas of Gabon were included. Biological and behavioral data were collected using an adapted version of the standardized WHO survey questionnaire.

The median age was 38[28–50] years. Tobacco consumption was more frequent in rural areas than in urban areas (26.1% vs 6.2%; *p* < 0.01). Men were more likely to be smokers than women, in both settings (aOR: 8.0[4.9-13.5], *p* < 0.01). Excessive alcohol consumption (19.4% vs 9.6%; *p* < 0.01) predominated in rural than in urban areas. Urban dwellers were less physically active than rural people (29.5% vs 16.3%; *p* < 0.01). In total, 79.9% of participants aged under 54 years had a high blood pressure (HBP) while 10.6% of the younger participants had pre-hypertension. Metabolic syndrome was higher in women (21.7% vs 10.0%; *p* < 0.01) than in men. Furthermore, 6.4% of men and 2.5% of women had a high risk of developing coronary heart diseases in the next 10 years (*p* = 0.03). Finally, 54.0% of the study population had three or four risk factors.

The prevalence rates of CMRFs were high in the study population. Disparities were observed according to urban and rural areas, gender and age groups. National prevention and healthcare strategies for cardiometabolic diseases in Gabon should take into account these observed differences.

## Introduction

Cardiovascular diseases (CVD) are the most common non-communicable disease (NCD). In 2019, CVD were responsible for an estimated 17.9 million deaths worldwide, of which more than three quarters occurred in low- and middle-income countries [1]. The increasing rate of deaths due to CVD remains a serious health concern. By 2030, 23 million people are expected to die from CVD, stroke and coronary heart diseases being the leading factors of these deaths. In sub-Saharan Africa (SSA), an increased prevalence rate of NCD, especially CVD, has been observed in the past 30 years. Ischemic heart diseases have now become a major leading cause of death in SSA, surpassing other major infectious diseases (malaria, tuberculosis and intestinal infections) [2].

Cardiometabolic diseases (CMD) are related to modifiable and non-modifiable risk factors (RF). Indeed, risk factors such as age, hypertension, diabetes, dyslipidemia, tobacco smoking, physical inactivity, high alcohol consumption, insufficient fruit/vegetable consumption and high fat intake are considered to be directly linked to the occurrence of CMD. High blood pressure (HBP) levels accelerate the risk of stroke and coronary heart disease. Likewise, people with diabetes are twice as likely to develop CVD. Physical inactivity is one of the lifestyle factors which promotes the onset of obesity, HBP, diabetes mellitus and an abnormal lipid profile. In SSA, HBP, type 2 diabetes, overweight and obesity were shown to be the most prevalent cardiometabolic risk factors (CMRF) [3].

Moreover, there has been a rapid increase of urbanization in SSA as well as significant population growth and aging in the past decades. Rapid and unplanned urbanization has been shown to contribute to the increase of CMD prevalence due to the increase of risk factors [4]. Indeed, unplanned urbanization is always characterized by the absence of spaces dedicated to physical activity. Furthermore, the urban environment provides an abundant market for the consumption of tobacco, alcohol and unhealthy food [5, 6].

Previous studies performed in high- and low-income countries highlighted disparities in the prevalence and types of CMRF in urban versus rural settings. For instance, in Cameroon, a higher prevalence of obesity (17.1%) and hyperglycemia (10.4%) has been reported in urban areas compared to rural areas [7]. In a study carried out in Burkina Faso, HBP was found in 24.8% and 15.4% of individuals living in urban and rural areas, respectively [8]. Furthermore, differences in the rates of risk factors according to gender have been described as well. Obesity and abdominal obesity are more often observed in women while men have a higher tobacco consumption [9, 10].

In Gabon, data on the burden of CMD and CMRF are scarce. Nevertheless, previous studies reported a non-negligible incidence of CVD (13.3%) and the prevalence of diabetes was estimated at 19.6% [11, 12]. Furthermore, according to the WHO global health statistics, one in five people is considered obese and in 2018, 17% of deaths were related to CVD [13]. In the same way, a high prevalence of HBP was described in young students [14]. It was recently shown that in SSA, the increased morbidity and mortality rates of CMD are a consequence of inefficient prevention programs and healthcare strategies [15]. The lack of data on CMD and their risk factors are key impediments to implement efficient control efforts. Recent data are required to develop awareness programs, to prioritize preventive measures and to adapt healthcare strategies according to urban and rural areas. As such, in order to better contextualize future prevention and healthcare strategies,

it is important to characterize the burden of CMRF according to sociodemographic factors (age, gender) and urbanization levels in Gabon.

Thus, the aim of this study was to estimate the prevalence rates of CMRF in urban and rural areas of Gabon.

## Material and methods

### Study sites

A prospective, cross-sectional and observational study was carried out from September 2020 to November 2021 in two urban areas (Melen and Libreville) and two rural areas (Bitam and Koula-Moutou) in Gabon. Melen is located in the fifth district of Libreville, the capital city. This district has 148,129 inhabitants. Koula-Moutou is located 586.9 km southeast from the capital, in the Ogooué-Lolo province. Bitam is located in the north of Gabon, 592 km from Libreville, in the Woleu-Ntem province. The Ogooué-Lolo and Woleu-Ntem provinces are the fourth and seventh most populated provinces in Gabon with 65,771 and 154,986 inhabitants, respectively. In these rural areas, the main activities of the population are agriculture, fishing and hunting.

The study was performed in Melen at the Centre de Recherche Biomédicale en Pathogènes Infectieux et Pathologies Associées (CREIPA), in Libreville at the Centre Hospitalier Universitaire de Libreville (CHUL), in Koula-Moutou at the Centre Hospitalier Régional Paul Moukambi (CHRPM), and in Bitam at the Centre Médical de Bifolossi.

### Study population

The study population was recruited through awareness campaigns in healthcare centers and in the community. Volunteers aged more than 18 years old, with negative malaria, HIV, hepatitis, tuberculosis tests, without known underlying disease, who had been residing in the four study sites for at least two years, and who gave their consent to participate were included. Pregnant women were not included.

### Sample size calculation

The sample size was calculated with the formula used to estimate a sample proportion: (n = z^2^ p(1-p)/e^2^) And the reported prevalence of cardiovascular disease in Gabon of 17%, in the absence of previous data on CMDRF [12]. The significance level was 5%. Considering a non-response rate of 10%, the minimum sample size was 239 included participants for rural and urban areas.

### Data collection and physical examination

Data were collected using an adapted version of the standardized WHO survey questionnaire by a team composed of experienced medical doctors, nurses and biologists [16]. The first phase of the study included a face to face interview with participants, during which they were asked about their socio-demographic data and their lifestyle characteristics such as tobacco and alcohol consumption, dietary habits (fruit, vegetable and salt consumption), physical activity. In addition, information on their past or current medication for HBP or diabetes was also recorded. In the second phase of the study, anthropometric measurements such as height, weight and waist circumference were taken. Height and weight were measured using a height gauge and an electronic

scale, respectively. Participants were weighed without shoes and wearing only light clothing. Measurements were recorded to the nearest 0.5 centimeter for height and 0.1 kg for weight. Body mass index (BMI) was calculated according to the following formula: weight (kg)/ height squared (m^2^). Waist circumference was measured (in cm) in a standing position, by measuring the midpoint between the lower margin of the last palpable rib and the top of the iliac crest, according to the WHO recommendations [16]. Blood pressure (BP) and heart rate were measured in the sitting position, on the left arm of each participant using an automatic blood pressure monitor (OMRON M3 Comfort, Kyoto, Japon) after 3 minutes of rest. Three measurements were performed 3 minutes apart and the average values of the two closest values of BP and heart beats were recorded.

### Biological analysis

After completing the questionnaire form and the physical examination, blood samples were collected in the morning from participants who had fasted for 12 hours. Blood glucose, total cholesterol (Total-C), HDL cholesterol (HDL-C), then LDL cholesterol (LDL-C) and triglyceride (TG) concentrations were measured using the PentraC200 analyzer (HORIBA ABX SAS, France).

### Quality procedure

Throughout the study, measurements were performed by two independent technicians. Participants with HBP and high fasting glucose levels, detected during screening, were seen 15 days later by the study team. If the same trends were observed, they were classified as having HBP or hyperglycemia. In the case of diagnosis confirmation, they were referred to the physician of the nearest health centre or case management. Data obtained were all comparable. Double data entry was carried out to minimize the risk of data entry errors.

### Definition of variables

The variables were defined based on WHO criteria [16]. Smokers were distributed into three groups: the first group included all tobacco users; the second group consisted of daily smokers (i.e. individuals who smoked at least one cigarette per day); the third group consisted of occasional smokers (i.e. people who smoke between 1 to 3 cigarettes per week). Passive smokers were people who had experienced passive smoking at home or at work during the 30 days prior to the day of the screening. Two groups were created for alcohol consumption: people who had consumed at least one alcoholic drink during the 12 last months, and those with excessive alcohol consumption, i.e. with more than 4 alcoholic drinks (12 g of ethanol) consumed per week. Different kinds of bottles or boxes were presented to the volunteers to allow the team to estimate the amount of drinking. After each volunteer reported the number of bottles or boxes he consumed, the physician estimated the standard quantity of drink which corresponded to 12 g of ethanol according to the most frequent type of drink available in the country (unpublished data). The number of fruit and/or vegetable intake per day was also assessed: an intake of less than 5 fruits or vegetables per day was considered as insufficient. Salt consumption was also evaluated and individuals were ranked according to their habits: occasional or excessive salting when cooking, adding salt before eating or usual consumption of salty additives with food.

Physical activity was estimated depending on the volunteers’ occupation, household activities, walking and physical activities during their free time. A physical activity <150 minutes per week was considered insufficient and a sedentary lifestyle was characterized by the fact of spending 12 hours of time sitting or lying down over the course of a day.

A metabolic syndrome was diagnosed according to the definition established by the International Diabetes Federation in 2005 [17]. The diagnosis was based on the combination of at least three of the five following RF: elevated triglycerides (>1.7 mmol/L); abdominal obesity (waist circumference ≥94 cm in men; ≥80 cm in women); systolic blood pressure ≥130 mmHg and/or diastolic blood pressure ≥85 mmHg or treated hypertension, fasting blood glucose ≥5.56 mmol/L or treated diabetes mellitus; low HDL cholesterol (HDL-C <1.03 mmol/L men, HDL-C <1.29 mmol/L women). Abnormal lipid levels were classified according to the Adult Treatment Panel III criteria as follows: high total cholesterol (Total-C ≥ 6.2 mmol/L), low HDL cholesterol (HDL-C <1 mmol/L); high LDL cholesterol (LDL-C ≥4.1 mmol/L) and high triglyceride levels (TG >2.26 mmol/L) [18].

HBP was defined as systolic blood pressure ≥130 mmHg and diastolic blood pressure ≥80 mmHg or when participants followed a treatment for HBP. Pre-hypertension was defined as systolic blood pressure between 120-129 mmHg and diastolic blood pressure between 70-79 mmHg [14]. An elevated heart rate amounted to >80 beat/min. High fasting blood glucose levels were defined by venous blood glucose levels ≥5.6 mmol/L after at least 12-hour fasting. Diabetes was defined when individuals followed treatment for diabetes mellitus. A BMI between 25 kg/m^2^ and 29 kg/m^2^ was considered as overweight, and obesity was defined as a BMI over 30 kg/m^2^. Abdominal obesity was defined for a waist circumference ≥94 cm in men and ≥80 cm in women.

The Framingham risk score for coronary heart disease was used to estimate the 10-year cardiovascular risk of the volunteers. The factors used in the calculation included age, gender, diabetes, smoking, SBP, DBP, Total-C and HDL-C. The cardiovascular risk score was categorized as low (<10%), intermediate (10%-19%), and high (≥ 20%) as recommended [19].

The following eleven biological and behavioral RF were used to assess the number of CMD risk factors in the age groups of the study population: HBP, hyperglycemia, diabetes, obesity, overweight, abdominal obesity, dyslipidemia, tobacco use, excessive alcohol consumption, low fruit/vegetable consumption, insufficient physical activity. Metabolic syndrome was considered as a group of RF.

### Ethical considerations

Ethical clearance and approval were obtained from the National Ethics Committee under the reference PROT N°002/2020/PR/SG/CNE. The study also received administrative authorization from the Ministry of Health (which is the national regulatory authority), as well as from the local administrators of each the study areas. All persons approached, including the authorities, were informed about the objectives of the study, the procedures, what was expected of the potential participants, the possible risks and benefits for them. For the illiterate participants, a local community member, previously trained by the research team, gave them information in the local language. Written and signed informed consent was obtained from each participant. A fingerprint was used as a signature for the illiterates who were accompanied by a witness who also signed the consent form. According to the National Ethics Committee requirement, the participants, aged below 21 years old, who agreed to participate signed a written assent form of consent and their parents or legal guardians provided a signed informed consent, after verification of their parental relationship. All participants and guardians were clearly informed about the study procedures, interest, potential risks and benefits, and their rights. Participants with abnormal clinical or biological data were conducted to the site health facility for appropriate investigation and management.

### Statistical analysis

Data were analyzed using Statistical Package for the Social Sciences (SPSS version 20) and Epi info (version 6). The data was further categorized according to areas, age group and gender. The normality of the data was checked by the Shapiro’s and Kolmogorov-Smirnov test for quantitative variables. Since the variables did not follow a normal distribution, medians were compared between groups using the Mann-Whitney U test and proportions were compared using a chi-squared test. Univariate logistic regression was performed to assess the unadjusted association between the studied variable and the presence of CMRF. All variables that were associated with the presence of at least one CMRF with a p<0.2 in the univariate analysis were included in the respective multivariable analysis. A multivariate multinomial logistic regression was used to determine the independent predictors of CMRF; we adjusted models for gender, areas of residence and age. Adjusted odds ratio (aOR) and cOR were reported at a 95% confidence interval (CI). All p-values are two-tailed, and p≤0.05 was considered statistically significant.

## Results

### Sociodemographic characteristics of the study population

In all, 1098 volunteers were approached and 978 were interviewed, 499 in urban areas and 479 in rural areas (Figure 1; Table 1). The results of the lipid measurements were available for 705 participants, 458 volunteers (154 men; 304 women) in urban areas and 247 volunteers (100 men; 147 women) in rural areas.

**Fig 1.**
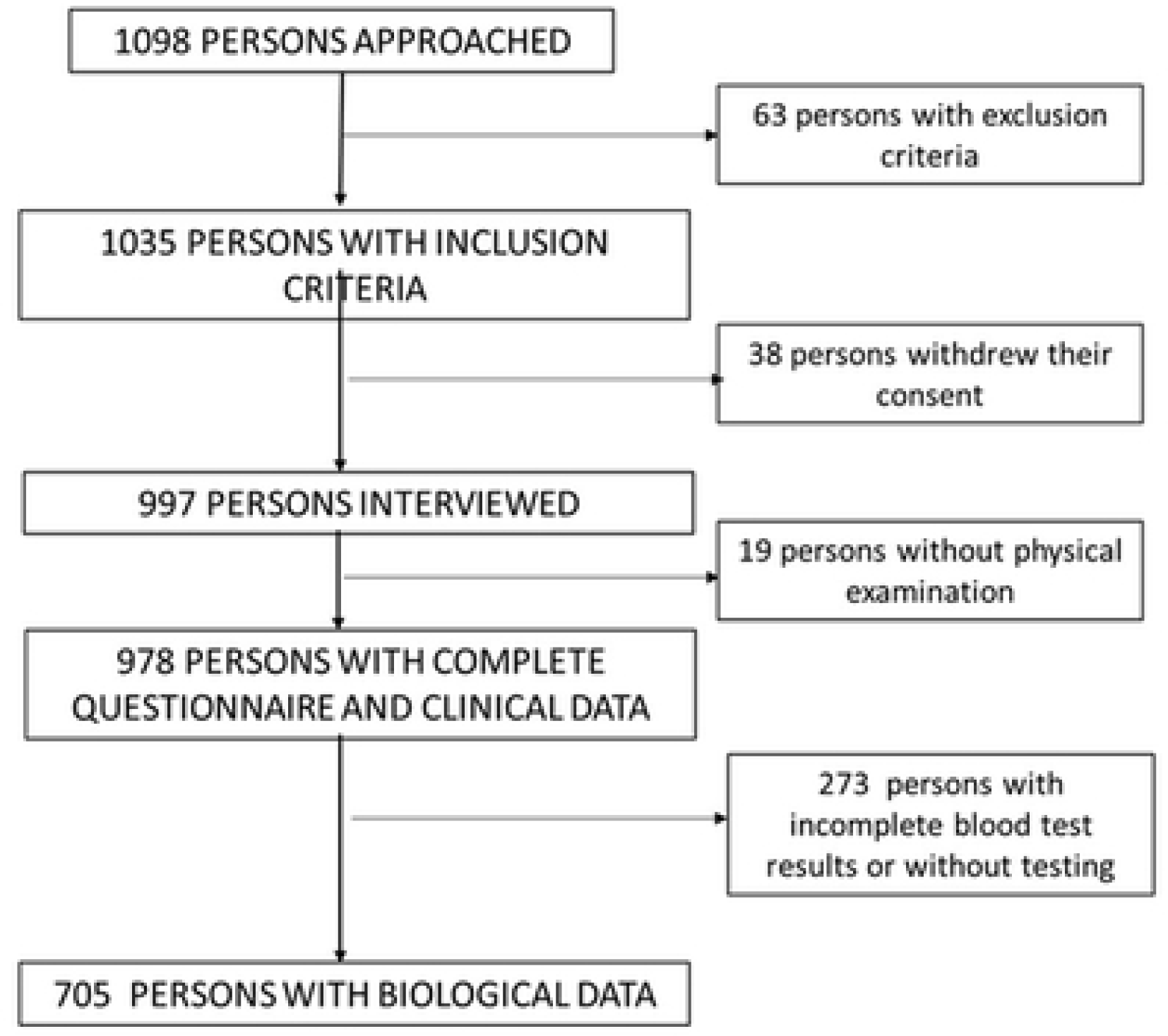
Study flow chart.

**Table 1.** Sociodemographic and socioeconomic characteristics of the study population.

| Characteristics | Total (N = 978) | Urban area (n = 499) | Rural area (n = 479) |
| --- | --- | --- | --- |
| **Gender n(%)** |  |  |  |
| Male | 405 (41.4) | 172 (34.5) | 233 (48.6) |
| Female | 573 (58.6) | 327 (65.5) | 246 (51.4) |
| **Age groups (years)** |  |  |  |
| 18-35 | 422 (43.1) | 255 (51.1) | 167 (34.9) |
| 36-53 | 395 (40.4) | 165 (33.1) | 230 (48.0) |
| >53 | 161 (16.5) | 79 (15.8) | 82 (17.1) |
| **Mean age (±SD)** | $39.5 \pm 12.6$ | $37.9 \pm 13.4$ | $41.0 \pm 11.6$ |
| **Marital status n(%)** |  |  |  |
| Unmarried | 379 (38.8) | 253 (50.7) | 126 (26.3) |
| Currently married | 261 (26.7) | 134 (26.9) | 127 (26.5) |
| Living with a partner | 283 (28.9) | 91 (18.2) | 192 (40.1) |
| Separated/Divorced/Widowed | 55 (5.6) | 21 (4.2) | 34 (7.1) |
| **Education level n(%)** |  |  |  |
| No formal education | 53 (5.4) | 23 (4.6) | 30 (6.3) |
| Primary school | 45 (4.6) | 22 (4.4) | 23 (4.8) |
| Middle school | 507 (51.9) | 157 (31.5) | 350 (73.0) |
| High school | 147 (15.0) | 93 (18.6) | 54 (11.3) |
| University education | 223 (22.8) | 202 (40.5) | 21 (4.4) |
| Refused to answer | 3 (0.3) | 2 (0.4) | 01 (0.2) |
| **Occupation n(%)** |  |  |  |
| Public sector worker | 117 (12.0) | 92 (18.4) | 25 (5.2) |
| Private sector employee | 137 (14.0) | 30 (6.0) | 107 (22.3) |
| Self-employed | 282 (28.8) | 70 (14.0) | 212 (44.3) |
| Worker/domestic worker | 72 (7.3) | 55 (11.0) | 17 (3.5) |
| Students | 166 (17.0) | 144 (28.9) | 22 (4.6) |
| Unemployed | 167 (17.1) | 80 (16.0) | 87 (18.2) |
| Retired | 35 (3.6) | 35 (3.6) | 09 (1.9) |
| Refused to answer | 02 (0.2) | 02 (0.2) | 00 (0.0) |

Women represented almost 60.0% of the study population. The majority of the participants were aged less than 54 years old (83.5%) and the median age was 38 years [28–50]. More than half (55.6%) and almost two thirds (66.9%) of the participants were in a relationship (married or living with a partner) and had at least a secondary school level education level (Table 1). Volunteers from urban areas were younger (37±13.4 vs 41± 11.6 years), more likely to be unmarried (50.7% vs 26.3%), and had a higher education level (40.5% vs 4.4%) than rural participants (*p* < 0.01) (Table 1).

### Prevalence of behavioral characteristics associated with cardiometabolic disease risk factors according to gender and area of residence

Overall, 156 (16.0%) participants smoked. Men were more likely to be smokers than women, both in rural (aOR: 8.0[4.9-13.5], *p* < 0.01) or urban settings (aOR: 4.0[1.8-8.4], *p* < 0.01) (Tables 2 and 4). Participants from rural areas were more frequently smokers (Table 2). Indeed, daily smokers (29.4% vs 19.8%, *p* < 0.01) were more frequent in rural areas than in urban areas. Overall, 24.7 % of the study population was exposed to passive smoking at work or at home.

**Table 2.** Behavioral characteristics associated with cardiometabolic disease risk factors according to study area and gender.

| Risk factors | Total | Urban |  | P-value<sup>a</sup> | Rural |  | P-value<sup>b</sup> | Urban | Rural | P-value<sup>c</sup> |
| --- | --- | --- | --- | --- | --- | --- | --- | --- | --- | --- |
|  |  | Men | Women |  | Men | Women |  |  |  |  |
| Tobacco consumption n(%) |  |  |  |  |  |  |  |  |  |  |
| All tobacco users | 156 (16.0) | 20 (11.6) | 11 (3.4) | < 0.001 | 103 (44.2) | 22 (8.9) | < 0.001 | 31 (6.2) | 125 (26.1) | < 0.001 |
| Daily smokers during previous 12 months | 106 (10.1) | 11 (6.4) | 03 (1.0) | < 0.001 | 84 (36.0) | 08 (3.3) | < 0.001 | 14 (2.8) | 92 (19.2) | < 0.001 |
| Occasional smokers | 50 (5.1) | 09 (5.2) | 08 (2.5) | 0.020 | 19 (8.2) | 14 (5.9) | 0.163 | 17 (3.4) | 33 (6.9) | 0.013 |
| Passive smoking during previous 30 days | 240 (24.5) | 44 (25.6) | 55 (16.8) | < 0.001 | 61 (26.2) | 80 (32.5) | 0.027 | 99 (19.8) | 141 (29.4) | < 0.001 |
| Alcohol consumption during previous 12 months | 640 (65.4) | 123 (71.5) | 206 (63.0) | 0.003 | 171 (73.4) | 140 (56.9) | < 0.001 | 329 (65.9) | 311 (64.9) | 0.741 |
| Excessive consumption during past 7 days | 141 (14.4) | 30 (17.5) | 18 (5.5) | < 0.001 | 70 (30.0) | 23 (9.4) | < 0.001 | 48 (9.6) | 93 (19.4) | < 0.001 |
| Fruits and vegetables intake |  |  |  |  |  |  |  |  |  |  |
| Number of fruits/days(m±SD) | 1.27 ± 1.2 | 1.20 ± 1.3 | 1.31 ± 1.3 | < 0.001 | 1.30 ± 1.2 | 1.23 ± 1.0 | 0.108 | 1.27 ± 1.3 | 1.27 ± 1.1 | 0.197 |
| Number of vegetables/day | 1.43 ± 0.8 | 1.28 ± 0.9 | 1.41 ± 0.8 | < 0.001 | 1.40 ± 0.8 | 1.59 ± 0.8 | 0.544 | 1.36 ± 0.8 | 1.50 ± 0.8 | 0.235 |
| Insufficient fruits/vegetables intake n(%) | 859 (87.8) | 150 (87.2) | 282 (86.2) | 0.641 | 206 (88.4) | 221 (89.8) | 0.469 | 432 (86.6) | 427 (89.1) | 0.218 |
| Salt consumption n(%) |  |  |  |  |  |  |  |  |  |  |
| Always/often adds additional salt during meals | 164 (16.8) | 30 (17.4) | 61 (18.7) | 0.621 | 36 (15.5) | 37 (15.0) | 0.857 | 91 (18.2) | 73 (15.2) | 0.209 |
| Usual consumption of salty additives in food | 320 (32.7) | 35 (20.3) | 86 (26.3) | 0.024 | 101 (43.3) | 98 (39.8) | 0.294 | 121 (24.2) | 199 (41.5) | < 0.001 |
| Physical activity n(%) |  |  |  |  |  |  |  |  |  |  |
| Low | 225 (23.0) | 41 (23.8) | 106 (32.4) | 0.002 | 35 (15.0) | 43 (17.5) | 0.288 | 147 (29.5) | 78 (16.3) | < 0.001 |
| Sedentary lifestyle | 88 (9.0) | 09 (5.2) | 36 (11.0) | < 0.001 | 13 (5.6) | 30 (12.2) | < 0.001 | 45 (9.0) | 43 (9.0) | 0.982 |

P-value<sup>a</sup>:Men-Women comparison in urban areas; P-value<sup>b</sup>:Men-Women comparison in rural areas; P-value<sup>c</sup>:urban-rural comparison.

Alcohol consumption was more prevalent than smoking in the general population, men being the most frequent users, whatever the study area (Table 2). Less than 15% of the study participants had excessive consumption, rural inhabitants and men were two-fold and 4-fold, respectively, at a higher risk of being excessive alcohol consumers (Tables 2 and 4).

Rural residents were more likely to eat food with additional salty additives than urban residents (41.5% vs 24.2%; *p* < 0.01) (Table 2). The same trend was observed for women living in urban cities (Table 2).

The mean fruit and vegetable consumption was 2.8 ± 2.3 /day. Only 12.2 % (n = 119/978) of volunteers consumed the recommended five or more fruits and/or vegetables per day (Table 2). Neither age nor area statistically influenced the quantity of fruit and vegetable intake, although women from urban settings had a higher fruit and vegetable consumption compared to men (Table 2). The rate of low physical activity was higher in urban areas (29.5% vs 16.3%; *p* < 0.01), and in women (26.0%, n=149/573 vs 18.8%, n=76/405; *p* <0.01) (Table 2).

### Prevalence of anthropometrics and biological cardiometabolic disease risk factors according to gender and area of residence

Almost, half of the study population had HBP (49.7%) (Table 3). HBP, diabetes, abdominal obesity, obesity, high LDL-C level, high Total-C level and metabolic syndrome were significantly more common in urban areas than in rural areas (Table 3).

**Table 3.**
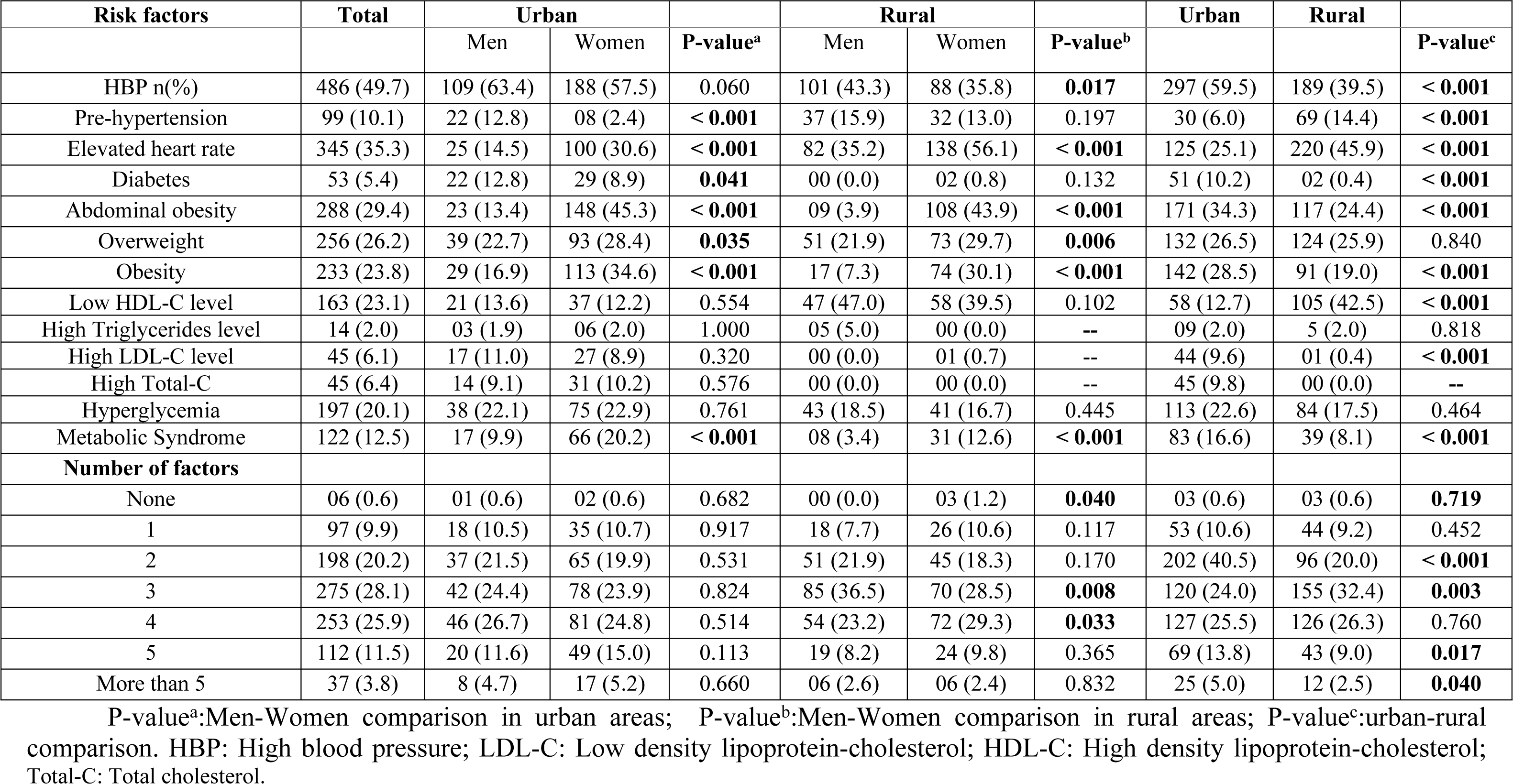
Biological risk factors, anthropometric data and number of cardiometabolic disease risk factors according to study area and gender.

The prevalence of HBP, including participants on medication for hypertension and pre-hypertension was 59.8% (Table 3). Among the respondents considered with HBP, 97(20%) were on medication. Men were more likely to have HBP or pre-hypertension than women (*p* < 0.01). However, while rural inhabitants had a lower prevalence of HBP, they were 1.5 times more at risk of having pre-hypertension (Table 4). Volunteers aged less than 36 years old were more likely to have pre-hypertension (10.6%) than older participants (3.9%) (*p* < 0.01). However, 36.5% of participants aged less than 36 years had HBP, compared to 62.2% of participants aged between 36 and 53 years old and 79.9% of participants who were older than 53 years old.

**Table 4.** Results of the logistic regression analysis for risk factors according to gender and study area.

| Variables | *Adjusted OR (95%CI) | P-value<sup>a</sup> | *Adjusted OR (95%CI) | P-value<sup>b</sup> |  | Crude OR | **Adjusted OR | P-value<sup>c</sup> |
| --- | --- | --- | --- | --- | --- | --- | --- | --- |
|  | Urban area |  | Rural area |  |  |  |  |  |
| **Tobacco users** | **Gender** |  | **Gender** |  | **Areas** |  |  |  |
| Male | 3.9 (1.82 – 8.39) | < 0.001 | 8.0 (4.86 – 13.45) | < 0.001 | Urban | Ref |  |  |
| Female | Ref |  | Ref |  | Rural | 5.33 (3.51-8.08 ) | 4.78 (3.1-7.3) | < 0.001 |
| Daily smokers |  |  |  |  |  |  |  |  |
|  |  |  |  |  | Urban | Ref |  |  |
|  |  |  |  |  | Rural | 8.23 (4.62-14.67) | 7.97 (4.38-14.53) | < 0.001 |
| Occasional smokers |  |  |  |  |  |  |  |  |
|  |  |  |  |  | Urban | Ref |  |  |
|  |  |  |  |  | Rural | 2.09 (1.15-3.81) | 2.43 (1.32-4.47) | 0.004 |
| Passive smoking |  |  |  |  |  |  |  |  |
|  |  |  |  |  | Urban | Ref |  |  |
|  |  |  |  |  | Rural | 1.68 (1.25-2.26) | 1.7 (1.27-2.31) | < 0.001 |
| **Excessive alcohol consumption** |  |  |  |  |  |  |  |  |
| Men | 3.7 (1.99 – 6.89) | < 0.001 | 4.1 (2.49 – 6.95) | < 0.001 | Urban | Ref |  |  |
| Women | Ref |  | Ref |  | Rural | 2.26(1.55-3.29) | 1.96 (1.33-2.88) | 0.0007 |
| **HBP** |  |  |  |  |  |  |  |  |
| Men | 1.6 (1.04 – 2.68) | 0.031 | 1.5 (1.03 - 2.30) | 0.032 | Urban | 2.25 (1.74-2.91) | 3.0 (2.23-4.12) | < 0.001 |
| Women | Ref |  | Ref |  | Rural | Ref |  |  |
| **Pre-hypertension** |  |  |  |  |  |  |  |  |
| Men | 8.7 (3,61 - 21.12) | < 0.001 | 1.5 (0.88 – 2.63) | 0.124 | Urban | 0.4 (0.24-0.59) | 0.6 (0.41-1.08) | 0.105 |
| Women | Ref |  | Ref |  | Rural | Ref |  |  |
| **Diabetes** |  |  |  |  |  |  |  |  |
| Men | 1.30 (0.68-2.49) | 0.421 |  |  | Urban | 27.15 (6.57-112.18) | 52.2 (12.3-221.3) | < 0.001 |
| Women | Ref |  |  |  | Rural | Ref |  |  |
| **Metabolic syndrome** |  |  |  |  |  |  |  |  |
| Men | Ref |  | Ref |  | Urban | 2.25 (1.52-3.36) | 2.46 (1.61-3.76) | < 0.001 |
| Women | 2.82 (1.55-5.12) | 0.0006 | 4.1 (1.83-9.14) | 0.0006 | Rural | Ref |  |  |
| **Overweight** |  |  |  |  |  |  |  |  |
| Men | Ref |  | Ref |  |  |  |  |  |
| Women | 2.3 (1.44 – 3.69) | < 0.001 | 2.5 (1.60 - 3.90) | < 0.001 |  |  |  |  |
| **Obesity** |  |  |  |  |  |  |  |  |
| Men | Ref |  | Ref |  | Urban | 1.69 (1.25-2.28) | 1.86 (1.31-2.63) | < 0.001 |
| Women | 4.2 (.54 – 7.14) | < 0.001 | 7.6 (4.22 - 13.74) | < 0.001 | Rural | Ref |  |  |
| **Abdominal obesity** |  |  |  |  |  |  |  |  |
| Men | Ref |  | Ref |  | Urban | 1.61 (1.22-2.13) | 1.55 (1.13-2.13) | 0.006 |
| Women | 7.2 (4.29 – 12.32) | < 0.001 | 20.4 (9.98 – 41.94) | < 0.001 | Rural | Ref |  |  |
| **Low physical activity** |  |  |  |  |  |  |  |  |
| Men | Ref |  | Ref |  | Urban | 2.14 (1.57-2.92) | 2.1(1.54-2.93) | < 0.001 |
| Women | 1.83 (1.8-2.83) | 0.006 | 1.33 (0.81-2.18) | 0.254 | Rural | Ref |  |  |
| **Sedentary lifestyle** |  |  |  |  |  |  |  |  |
| Men | Ref |  | Ref |  |  |  |  |  |
| Women | 3.04 (1.39-6.65) | 0.005 | 2.57 (1.28-5.15) | 0.007 |  |  |  |  |
| **Usual consumption of salty additives in food** |  |  |  |  |  |  |  |  |
|  |  |  |  |  | Urban | Ref |  |  |
|  |  |  |  |  | Rural | 2.22 (1.68-2.91) | 2.39 (1.8-3.1) | < 0.001 |
| Low HDL –c level |  |  |  |  | Urban | 0.5 (0.33-0.66) | 0.2 (0.13 – 0.29) | < 0.001 |
|  |  |  |  |  | Rural | Ref |  |  |
| High LDL-c level |  |  |  |  | Urban | 46.2 (6.34 - 336.8) | 35.0 (4.7-257.4) | < 0.001 |
|  |  |  |  |  | Rural | Ref |  |  |
| 2 RF |  |  |  |  | Urban | 2.71 (2.03 - 3.61) | 1.49 (0.2 -7.73) | 0.633 |
|  |  |  |  |  | Rural | Ref |  |  |
| 3 RF |  |  |  |  |  |  |  |  |
| Men |  |  | 1.13 (0.34-3.76) | 0.839 | Urban | 0.66 (0.5 - 0.87) | 1.20 (0.23 - 6.22) | 0.824 |
| Women |  |  | Ref |  | Rural | Ref |  |  |
| 5 RF |  |  |  |  | Urban | 1.62 (1.08-2.4) | 2.66 (0.49 - 14.20) | 0.252 |
|  |  |  |  |  | Rural | Ref |  |  |
|  |  |  |  |  | **Number of RF** |  |  |  |
|  |  |  |  |  | **One RF** |  |  |  |
|  |  |  |  |  | 18-35 | Ref |  |  |
|  |  |  |  |  | 36-53 | 0.44 (0.27-0.70) | 0.46 (0.16-1.33) | 0.154 |
|  |  |  |  |  | > 53 | 0.20 (0.07-0.50) | 0.10 (0.02-0.40) | 0.001 |
|  |  |  |  |  | **2 RF** |  |  |  |
|  |  |  |  |  | 18-35 | Ref |  |  |
|  |  |  |  |  | 36-53 | 0.55 (0.39-0.77) | 0.38 (0.14-1.01) | 0.055 |
|  |  |  |  |  | > 53 | 0.42 (0.25-0.70) | 0.17 (0.05-0.49) | 0.001 |
|  |  |  |  |  | **3 RF** |  |  |  |
|  |  |  |  |  | 18-35 | Ref |  |  |
|  |  |  |  |  | 36-53 | 1.09 (0.81-1.46) | 0.59 (0.22-1.51) | 0.274 |
|  |  |  |  |  | > 53 | 0.9 (0.60-1.35) | 0.29 (0.10-0.79) | 0.017 |
|  |  |  |  |  | **4 RF** |  |  |  |
|  |  |  |  |  | 18-35 | Ref |  |  |
|  |  |  |  |  | 36-53 | 1.59 (1.16 -2.18) | 0.86 (0.33-2.24) | 0.761 |
|  |  |  |  |  | > 53 | 1.85 (1.26 -2.73) | 0.59 (0.21-1.63) | 0.316 |
|  |  |  |  |  | **5 RF** |  |  |  |
|  |  |  |  |  | 18-35 | Ref |  |  |
|  |  |  |  |  | 36-53 | 1.82 (1.14-2.90) | 0.84 (0.30-2.32) | 0.743 |
|  |  |  |  |  | > 53 | 2.36 (1.37-4.07) | 0.55 (0.18-1.62) | 0.281 |
|  |  |  |  |  | **> 5 RF** |  |  |  |
|  |  |  |  |  | 18-35 | Ref |  |  |
|  |  |  |  |  | 36-53 | 2.13 (0.90-5.04) |  | 0.083 |
|  |  |  |  |  | > 53 | 4.25 (1.73-10.46) |  | 0.001 |

\*AOR adjusted for age/ \*\*AOR adjusted for age and gender / For the number of risk factors, OR was adjusted according to gender, the education level and occupation. P-value<sup>a</sup>:Men-Women comparison in urban areas; P-value<sup>b</sup>:Men-Women comparison in rural areas; P-value<sup>c</sup>:urban-rural areas comparison. RF: Risk factor; Ref: Reference; HBP: High blood pressure; LDL-C: Low density lipoprotein-cholesterol; HDL-C: High density lipoprotein-cholesterol.

The mean waist circumference and BMI were, respectively, 84.82 ± 15.53 cm and 26.50 ± 6.64 kg/m^2^ in the overall study population. Abdominal obesity was found in more than 20% of the participants, while the overall prevalence of overweight and obesity was 50.0% (Table 3). Living in urban areas (aOR: 1.89, 95%CI [1.31-2.63], *p* < 0.01) and female gender (aOR: 4.2, 95%CI [3.54 – 7.14], *p* < 0.01) were associated with obesity (Table 4). According to age, a higher prevalence of obesity was seen in adults aged 36-53 years old (34.5%) compared to the oldest (29.8%) and the youngest (16.4%; *p* < 0.01). The prevalence rate of overweight was comparable between the three age groups, ranging from 25.0% to 29.1%.

The mean (±SD) of Total-C, LDL-c, HDL-C, TG levels were 4.89 (±1.29) mmol/L, 2.43 (±1.07) mmol/L, 1.36 (± 0.52) mmol/L and 0.82 (± 0.52) mmol/L, respectively. Low HDL-C (23.1%) was the most prevalent dyslipidemia, while the rates of other markers were below 7.0% (Table 3). There was higher rate of low HDL-C in rural areas. High LDL-C and hypercholesterolemia were more frequent in inhabitants from urban sites. However, no difference was observed between men and women for high Total-C levels (5.5% in men vs 6.3% in women; *p* = 0.47), and for high LDL-C (6.7% in men vs 6.2% in women; *p* = 0.92) (Table 3).

Our results show a higher trend towards hypertriglyceridemia (3.2% in men vs 1.3% in women; *p* = 0.10) and low HDL-C (26.8% in men vs 21.1% in women; *p* = 0.09) in men compared to women. The mean Total-c rate was 6.9 (± 0.7) mmol/L in the group of volunteers with a high Total-C, the mean HDL-C rate was 0.69 (± 0.23) mmol/L in participants with low HDL-C, the mean LDL-C rate was 4.91 (± 0.93) mmol/L in participants with high LDL-C, and finally, the mean TG was 3.26 (± 0.49) mmol/L in the group of volunteers with hypertriglyceridemia.

The overall prevalence of diabetes was 5.4% and the prevalence rate of high fasting blood glucose levels was almost 4-fold higher (Table 3). The hyperglycemia prevalence rate was comparable in urban and rural areas, while 51 out of the 53 volunteers on diabetes medication lived in urban sites (p < 0.01). The burden of both abnormalities was not statistically different according to gender. The overall prevalence of metabolic syndrome was higher in women (21.7%) than in men (10.0%; *p* < 0.01) (Table 3).

### Framingham cardiovascular risk score according to urban and rural areas

The Framingham risk score for coronary heart disease was assessed only in the group of the 505 participants aged over 30 years old (296 in urban areas and 209 in rural areas). Overall, 30 (3.4%) participants had a high score, and among these 18 (6.1%) lived in urban settings while only 2 (1.0) resided in rural areas (*p* < 0.01). The rates of people with a moderate score were comparable in the two areas (12.5% in urban area vs 11.0% in rural area, *p* = 0,61). The prevalence of a high risk score was significantly lower in men (6.4% in women vs 2.5% in men; *p=*0.03), and in older participants (13.7% vs 1.4% in those aged between 36-53 years old; *p <* 0.01).

### Number of cardiometabolic disease risk factors according to gender, age group and area of residence

Overall, 972 (99.4%) participants had at least one modifiable CMD risk factor. Participants frequently had three or four RF (54.0%) and those with the highest number of risk factors resided in urban areas (Table 3).

The number of CMRF per individual significantly varied according to age (*p* < 0.01) (Fig 1). The majority of healthy volunteers aged under 36 years old (70.9%; n = 299/422) had three RF or less; two thirds of the oldest participants (60.3%, n = 97/161) had four RF or more, while the middle-aged volunteers had more frequently (60.0%, n = 237/395) three to four CMRF (Fig 2).

**Fig 2.**
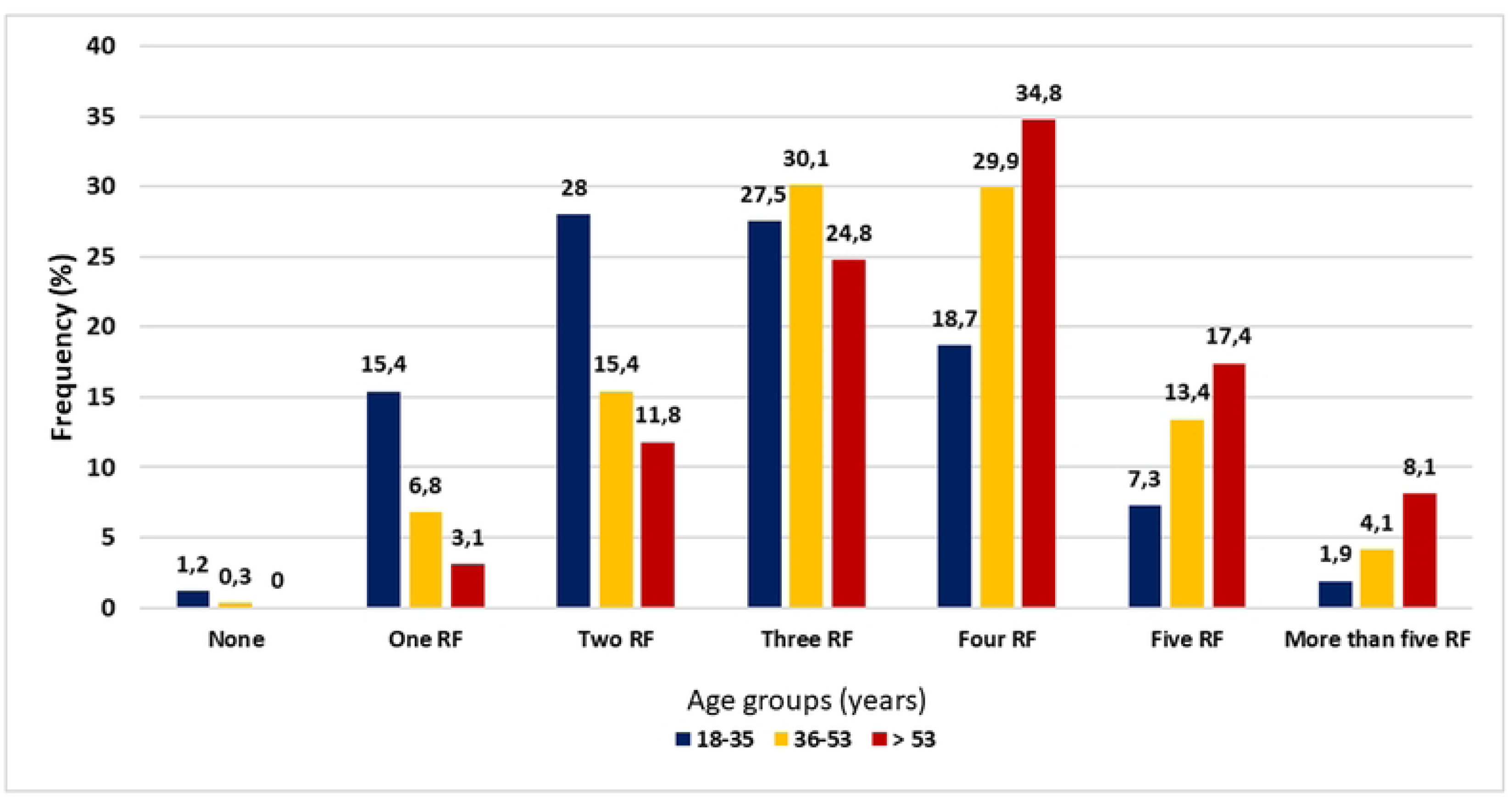
Number of cardiometabolic disease risk factors according to age groups.

## Discussion

The burden of NCD is rapidly growing worldwide, including in SSA where populations also suffer from the burden of infectious diseases. Public health efforts mainly focus on reducing the burden of the three main infectious diseases (malaria, HIV/AIDS and tuberculosis) and neglected tropical diseases (NTDs). However, reports on the increasing death rate associated with CVD are on the rise for SSA [12, 20]. In addition, data from Central Africa are scarce, although the living conditions of local populations are not significantly different from other low-middle-income-countries in SSA. This lack of reliable data is an obstacle for (i) the consideration of NCD as serious public health issue, and (ii) for the design and planning of efficient preventive strategies tailored to the population, in order to reduce the silent and neglected impact of NCD.

This study shows a considerable high prevalence of individual NCD risk factors in people living in urban and rural settings in Gabon.

To date, there was no recorded data on the prevalence of daily tobacco use and harmful alcohol consumption in Gabon, although these behaviors are noticed among young adults. The prevalence of daily tobacco use was higher than that observed in young Gabonese students (19.2% vs 14%), in Kenyan adults (13.5%) and in global reports from SSA [14, 21, 22]. The fact that men are more likely to be smokers than women is currently reported worldwide [9, 23]. However, an unusual higher prevalence of daily tobacco users in rural areas (19.2%) compared to urban settings (2.8%) was observed. In SSA countries, the smoking prevalence rate was found to be 40% in urban areas versus 20% in rural areas [24]. Approximatively, 25% of the study population were exposed to passive smoking at work or home. Although Gabon has adopted and designed preventive strategies after implementing the anti-smoking law in 2013, it seems that awareness campaigns are insufficient in the country and have difficulty reaching rural communities with lower school attendance. This study emphasizes the need to continue data collection and to consider gender and sociodemographic factors, in order to tailor preventive strategies to target populations. Rural communities must be better informed and included in the preventive programs for NCD.

Smoking and alcohol consumption are two RF for CMD which often occur together. Excessive alcohol consumption was more prevalent in men and in rural settings as observed elsewhere [9, 10]. Alcohol and tobacco consumption in women is traditionally considered as abnormal or incorrect (immoral) in Gabon, while it is more accepted for men, this could explain the difference highlighted according to gender. It is important to note that this gender difference is only observed for the excessive consumption of tobacco or alcohol, whatever the level of urbanization. Surprisingly, smoking and alcohol consumption were more often reported by rural inhabitants. Alcohol shops are easily accessible in urban settings, but local production is frequent in rural settings. Each family can have its own palm plantation and sugar cane fields as well as homemade stills, and therefore have the possibility of producing its own palm wine or cane sugar liqueur whose alcohol level can exceed 30%. The two drinks, which are very popular in local communities, are very often consumed without moderation.

An unhealthy diet and insufficient physical activity are common in the study population, with prevalence rates comparable to findings from other African countries. Indeed, the prevalence of low fruit and vegetable intake is in line with reports from Tanzania (82%) and Ethiopia (95%) [25, 26]. There was no difference according to gender or the geographic location. This could be due to the population’s dietary habits and their similar access to various fruits and vegetables. Most participants in this study (74%) did not have a sufficient and regular income which would allow them to buy and consume several fruits and vegetables on a daily basis. In addition, the eating habits of the population depend on the seasonal availability of foods and also on the geographic area. Although agriculture is the main activity in rural areas and these are the main supplier of local fruits and vegetables, the availability of these is seasonal and there is only very little variety and quantity: certain fruits are found in a region at a given time, and not necessarily in another. In addition, the average Gabonese meal consists of a starchy food and meat or fish for lunch, which is the main meal of the day. The consumption of fruit for snacks or dessert is only possible for people who have sufficient income to buy expensive imported fruits and vegetables every day. The questionnaire used during the interviews did not take into account the local context to gather data on the availability and specificities of local foods; future studies should adapt the questions accordingly. Gabonese government policies should promote healthy eating habits, for example by making healthy foods available at reduced prices in urban areas and by providing agricultural land and mechanical tools dedicated to agriculture in rural areas.

The prevalence rate of high salt intake in cooking and dietary habits was not negligible. It reached 42% in total when the consumption of processed food with salt was combined with the addition of salt during meals. The higher prevalence observed in rural areas could be explained by the absence of preventive information about the risk of CVD when eating more than 5g of salt per day, but also by the fact that cardiologists, who usually provide such education and counselling, are lacking in rural areas.

The overall prevalence of low physical activity is 22% in SSA, which is in line with the results in our present study (23%). The higher proportion of women and urban participants with insufficient physical activity seems to be common in low-and middle-income countries [27]. The sedentary lifestyle (at home and at work) in urban cities and the fact that men do more physical work compared to women might partly be in cause.

The prevalence rate of overweight and obesity in the study population is cause for concern as it amounts to 50%. This is very high compared to reports from other SSA countries such as Mozambique (30%) and Zambia (24.4%) [28, 29]. The prevalence of central obesity (29%) was however comparable to data from these countries. Obesity is the consequence of energy imbalance between the type and quantity of food consumed and insufficient physical activity. The predominance of both CMRF among women and the urban population of this study justify the observed higher predominance of overweight/obesity and central obesity in these two groups. Our results are in accordance with other data in African countries where the prevalence rate of overweight/obesity has doubled within the last two decades, with the highest burden found in women and urban inhabitants [30, 31]. The westernized and sedentary lifestyle of urban populations of low-middle-income-countries results in infrequent physical inactivity during free time, more time spent in offices in front of computers, increased use of public transportation, and the increased consumption of processed foods and junk foods, which are all causal factors for overweight/obesity. In Libreville, the work is carried out in a continuous day with a break of less than an hour. Workers therefore do not have the possibility of returning home to have a balanced lunch, the majority having lunch in fast food outlets where fried foods and meals rich in oil and salt are most frequently available. This poor lifestyle may explain the higher prevalence of obesity in the group of patients aged 35 to 53 years old, which is the most professionally active in our study population.

Hypertension is the leading CMRF in Africa, its prevalence is the highest in the world, ranging from 17% to 70% [32]. Almost half (49.7%) of the interviewed persons in our study had a high blood pressure, which is consistent with a previous report from the main hospital in Libreville, but considerably higher compared to studies from Oman (33) [33, 34]. Although the differences could be due to the cross-sectional study design and other parameters such as the stress of the participants during the physical examination, these findings are a matter of grave concern for physicians, health workers, and policy makers. Indeed, the prevalence of participants on medication is still high (20%). Our findings also suggest that more than 25% of the population were not aware of their health status or were undiagnosed, and thus would remain untreated, increasing the risk of ischemic attack. There is an urgent need for preventive efforts, better diagnostics, high treatment coverage, and awareness campaigns on the causes and consequences hypertension. The prevalence of HBP was lower in rural settings in this study, it reached 39% in areas where it is known that awareness campaign and annual systematic screening and treatment are not often accessible. A recent study conducted in Libreville revealed an HBP prevalence rate of nearly 20% among adolescents [14]. Altogether, these results suggest that HBP is a major public health problem in Gabon and that there is an urgent need to implement control and prevention strategies. These findings also highlight the need for an integrated approach of NCD for prevention in urban and remote rural areas.

The risk of developing a CVD increases with high fasting blood glucose levels; more than 20 million African individuals live with diabetes [35]. Data on the prevalence of diabetes in Central Africa are sparse. The prevalence rate of diabetes was 5.4% in this study, which is in line with studies from Cameroon (5.6%), Burkina Faso (5.2%) [36, 37]. However, not surprisingly 97% of the diabetic participants lived in urban areas, and as such the prevalence rate was higher (10.2%) than in rural areas. The prevalence of hyperglycemia was much higher than that of diabetes, and it was comparable in urban and rural settings. Beyond the deaths directly attributable to diabetes, hyperglycemia is responsible for a heavy burden of mortality [38]. It is therefore urgent to implement a strategy to treat patients in the pre-diabetes stage in order to avoid a progression of the disease to diabetes and its medical and economic consequences.

Effective detection and treatment of dyslipidemia are recommended to reduce cardiovascular diseases in Africa. The burden of these biological risk factors is increasing in SSA [20]. Low HDL-C level was the most frequent detected dyslipidemia (23.1%), which is consistent with data from SSA (20-40%) [39]. High Total-C (6.4%) and hypertriglyceridemia (6.3%) prevalence rates were lower than the majority reported data from other SSA countries [39, 40]. However, the concerning prevalence rate of the metabolic syndrome (12.5%), which is a precursor of CVD and diabetes, is in line with rates from SSA (16%) [41]. Women and urban populations are the most concerned. This is not surprising because related behavioral risk factors were also found to predominate in these groups. Pre-HBP (14.4% vs 6.0%), elevated heart rate (45.9% vs 25.1%) and low HDL-C (42.5% vs 12.7%) levels were predominant in rural areas. These results could reflect an ongoing epidemiologic transition, in rural areas.

Less than 1% of the apparently healthy study participants had no CMRF. More than 80% had at least two NCD risk factors, although persons living in urban sites (18.8%) were more likely to have more than three RF than rural inhabitants (11.5%). In Kenya, the majority (75%) of adults screened had at least four RF [42]. As observed elsewhere, increasing age and urban residence were associated with a higher number of CMRF [28].

According to the Framingham risk score, people at a high risk of suffering from a cardiovascular event in next 10 years were more likely to live in urban areas than in rural areas (6.08% vs 0.95%), as reported in Myanmar [9]. These results are not surprising as the variables considered in the calculation of the Framingham risk score were found in high prevalence rates in urban sites.

Urban inhabitants had the most risk factors and older people had a higher number of RF. It has been suggested that the risk of cardiovascular events increase with particular combinations of risk factors as well as their total number [43, 44]. As an example, hypertension associated with the elderly may be combined with several others RF. Thus, strategies to reduce RF in the population should be adapted to target the population according to the areas.

This is the first study which assessed CMRF both in urban and rural areas of Gabon, were no data were previously recorded. A standardized approach was used, making the generated data comparable to other countries. Data was collected by well-trained team members, and the presence of a field worker living in each of the study area and speaking the native language, facilitated exchanges with the populations in rural localities. However, although the Framingham risk equation was used to assess the 10-year risk score of cardiovascular events, it has not been validated in African populations. The development and validation of a region-specific risk estimation equation is much needed. The recorded data were self-reported through face-to face interviews on personal lifestyle. Thus, the participants may tend to say what is acceptable or correct instead of being honest. As a consequence, alcohol and tobacco consumption could be under-reported. Moreover, the measurement of salt in urine was not performed to assess the reliability of the volunteers estimates of their salting habits. Lastly, this study was cross-sectional and did not assess environmental and genetic risk factors. Despite these limitations, the data in this study provide useful baseline information on the burden of CMRF and could be used by policy makers and control programs.

## Conclusion

This study provides essential information on CND indicators in Gabon. The prevalence of modifiable CMD risk factors was high both in urban and rural study sites. Gender differences in risk factor prevalence was observed. The high prevalence of multiple RF per individual indicates the urgent need to implement effective preventive programs as well as the detection, treatment and prevention of related cardiovascular and metabolic diseases. Other surveys with larger sample populations and specific groups such as youth, elderly, pregnant women and HIV-infected persons should also be performed.

## Data Availability

The data will be sent on demand

None

## Acknowledgements

The authors would like to thank the EDCTP2 program for funding this work. Many thanks to the Département de Parasitologie-Mycologie, Université des Sciences de la Santé, Owendo Gabon, as well as the staff of the Centre de Recherche Biomédicale en Pathogènes Infectieux et Pathologies Associées (CREIPA) of Melen for their help in carrying out this work.

